# The molecular basis for prognosis of isoniazid resistance in *Mycobacterium tuberculosis*

**DOI:** 10.1101/2024.09.21.24314105

**Authors:** Siavash J. Valafar, Aram A. Valafar, Wael Elmaraachli

## Abstract

Tuberculosis (TB), a disease that kills 1.5 million people every year, is a major global public health concern. The emergence of drug resistance in *M. tuberculosis*, the obligate pathogen of TB is a major challenge. The emergence of resistance seems to follow an order that might be exploited for novel therapeutic strategies. In most cases resistance to isoniazid (INH) emerges first, followed by rifampicin, then either pyrazinamide or ethambutol, and finally followed by resistance to second-line drugs. For this reason, it is thought that prevention of emergence of INH resistance may help the prevention of resistance to other drugs. In this manuscript we present the prognostic potential of specific mutations in predicting the emergence of the three most common canonical INH resistance (katG315, inhA-15, and inhA-8) with the hope that majority of resistance cases can be predicted and avoided. Here we present evidence that resistance to INH occurs in steps that in most cases follow specific evolutionary trajectory. Identifying these steps can therefore be used to predict and avoid the most common INH resistance mechanism. In our approach, we used genomic and phenotypic data from over 16,000 samples collected by two large databases, the TB Portals and the CRyPTIC consortium. We used classical sensitivity and specificity values as well as a deep learning neural models to identify promising predictive mutations using TB Portals data. We then tested the prognostic potential of the identified mutations using the CRyPTIC consortium data. Here we report two mutations (*Rv1258c* 581 indel & *mshA* A187V) as those carrying the highest potential for predicting the emergence of the three canonical mutations (accuracy of 73% and specificity of 96%). Our results point to a stepwise evolutionary trajectory toward the emergence of the three canonical mutations. Furthermore, the high negative predictive values provide an opportunity for clinicians to continue using INH in new regiments designed for nonresponsive patients whose samples do not contain the two precursor mutations. Finally, we present testable hypotheses describing the role of the precursor mutations in emergence of the three canonical mutations and the predicted trajectories. Mutagenesis experiments can confirm these hypotheses. Additional time course samples and analysis will undoubtedly uncover additional prognostic markers for other trajectories toward high-level INH resistance.

## 1. Introduction

*Mycobacterium tuberculosis* (*M. tuberculosis*), the causative agent of the tuberculosis (TB) disease, infects millions annually.^1^ Annually, 10.5 million people develop the active form of the disease for the first time, globally.^1^ TB is commonly treated by the standard regimen recommended by the World Health Organization (WHO) which includes the combination of the four “first-line” drugs ethambutol (EMB), isoniazid (INH), rifampicin (RIF), and pyrazinamide (PZA).^2^ Unfortunately, the emergence of antibiotic resistance is a major challenge in global TB control.^1^ The WHO estimates 1.3 million people developed isoniazid resistant (INH^R^) TB, globally.^1^ This included an estimated 410,000 cases of multidrug resistant tuberculosis (MDR-TB) where the bacterial strains are resistant to both RIF and INH.^1^ Treatment of MDR-TB is challenging and estimated to succeed only in 52% of cases (without new and repurposed drugs [NRDs]).^3^ Yet more complex is the treatment of extensively drug resistant tuberculosis (XDR-TB) cases where the strains are MDR-TB but also resistant to any fluoroquinolone and at least one additional Group A drug. Treatment of XDR-TB cases is only successful in less than 25% of cases without NRDs (bedaquiline, delamanid, pretomanid, clofazimine, and linezolid). While NRDs have offered renewed hope and significantly improved the success rate of MDR-TB and XDR-TB treatment (between 60%^4^ and 89%^3^ for MDR and 48.6%^5^-90%^6^ for XDR-TB in some trials with the six-month BPaL or BPaLM regimens), the cost of these drugs, their availability, and increasing incidence of resistance to them in programmatic application globally is a distinct concern.

Emergence of antibiotic resistance generally follow an order where INH resistance emerges first, followed by RIF resistance, followed by resistance to PZA or EMB, and finally to second line drugs.^7–12^ It has been hypothesized that prevention of emergence of INH resistance increases the likelihood of sterilization and reduces the likelihood of progression to MDR-TB^13–16^ through timely increased INH dosing or replacement with another drug alternative. Three mutations in the *M. tuberculosis* genome are responsible for an estimated 81.4% of all reported INH^R^ cases globally.^17^ By far the most common INH resistance-causing mutation is in codon 315 of the catalase peroxidase gene *katG* (*katG*315), followed by the -15 and -8 mutations in the promoter of the operon that includes the *inhA* gene (*inhA*-15, *inhA*-8).^17^ In this article we explore the potential of using genomic markers in *M. tuberculosis* for prognostic purposes. Specifically, we ask whether any genomic markers in *M. tuberculosis* can predict the emergence of these three INH resistance-casing mutations. We used a deep learning artificial intelligence (DL-AI) model to identify two markers for this prognostic purpose. We then used the Cryptic consortium genome collection to estimate the prognostic value of the two mutations.

## 2. Methods

### Data

In total 11,747 sets of whole genome sequencing (WGS) and associated INH drug susceptibility testing (DST) results were included in this study from two publicly available sources:

a. *TB Portal*: The first set included 4,131 clinical isolates from TB Portals^18^ for training the DL-AI model. Of these, we selected for inclusion 1,931 genomes and their associated INH DST collected from 453 patients. The selection criteria for these were that each patient had to have at least two time-course samples that 1) one sample was from prior to their treatment start (baseline sample) and at least one was from after treatment start, 2) each had associated INH DST data, and 3) had WGS data available for the nine INH-resistance associated genes.
b. *CRyPTIC Consortium*: The second dataset included 12,289 clinical isolates from the CRyPTIC consortium^19^ which was used for the assessment of the prognostic sensitivity of the DL-AI identified markers. Of the total of 12,289 samples stored in the CRyPTIC database, we included 9816 genomes from 919 patients. The selection criteria for these were the same as those for the TB Portals samples.

### Patient Categorization

Patient groups from both sets were divided into three categories:

a. EMERGED: group of patients for whom the emergence of at least one of the three canonical mutations (*katG*315, *inhA*-15, *inhA*-8) were observed during their treatment.
b. INITIAL RESISTANCE: group of patients for whom at least one of the canonical mutations was observed in the baseline sample.
c. NOT EMERGED: group of patients for whom the three canonical mutations were never observed in any of their samples during their treatment.

### Selected genes

From our previous systematic review^17^ and a literature search, we identified nine genes associated with INH resistance: *katG, inhA, ndhA, fabG1, ahpC, Rv1258c, mshA, Rv2752c*, and *ndh*. In this exercise some genes had to be dismissed from this study due to their extensive Illumina “blind spots”.^20^ Blind spots are loci in the *Mycobacterium tuberculosis* genome that are not sequenced properly by Illumina platforms because of various reasons.^20^ For instance 170 (11.8%) positions in the gene *iniB* are Illumina blind spots.^20^ Because both data sets that we used exclusively used Illumina sequencing, we did not include this gene in our study. While this is not a comprehensive list since genes such as *kasA*^21^ and *furA*^21^ are excluded, the nine genes considered in this study represent the most sited genes in the literature as playing a role in INH resistance.

### Deep Learning (DL)

In total the genomic and phenotypic data from 453 patients from TB Portal to train the DL model. The goal of this exercise was to identify the mutations that carried the highest prognostic power for prediction of emergence of the three canonical mutations (*katG*315, *inhA*-15, *inhA*-8) independent from the CRyPTIC data. For each patient, two sets of samples were prepared.

#### Input data

Mutations in the listed nine genes from the baseline sample (TB Portals samples) of each patient were provided to the models as binary input data. In total 49 mutations (Supplementary Table 1) were identified in these genes across all patients (Supplementary Table ST1). For each patient a binary vector of presence/absence vector (length 49) was generated based on the patient’s baseline genomic data.

#### Desired target data

The last sample of the patient was evaluated for the presence of the three canonical mutations. For patients whose follow up samples (any sample other the baseline sample) contained any of the canonical mutations, a desired target value of 1 was designated, indicating the emergence of the canonical mutations. If none of the canonical mutations were observed for any of the time-course samples of the patient, a value of 0 was designated as the desired target.

#### Model topology, training, and experimental design

In this study, we only explored the prognostic value of up to two mutations. As such, a Deep Neural (DN) model was trained for each of the 49 mutations individually, and one was trained for any combination of two mutations from the set of 49 (1,176 combinations). As a result, the total number of trained models was _*49*_*C*_*1*_ + _*49*_*C*_*2*_ = 1,225.

All models had a three-layer topology with two hidden layers and one output layer. Size of the three layers for the single-mutation models were optimized empirically to be 3-2-1. While layer optimization for the two-mutation models resulted in 5-3-1 networks. For each model a 10-fold cross-validation process was used to evaluate the performance of the model. We split the 453 patients into a training set (including cross-validation) of 250 randomly selected patients and a testing set of 203 patients. The random selection was limited to patients within each of the three categories of EMERGED, INITIAL RESISTANCE, and NOT EMERGED sets to ensure that each of these categories were properly represented in both the training and the testing sets. The trained models were then used to predict the emergence of any of the three canonical mutations in the testing set. For these experiments the Deep Learning/Statistics and Machine Learning toolboxes of MATLAB version R2023a were used.

#### Validation of the DL results

In order to validate the results of the DL models, we used 9816 genomes from 919 patients from the CRyPTIC consortium^19^ as an independent validation set.

### Sensitivity, Specificity, Positive Predictive Value (PPV), and Negative Predictive Value (NPV) assessment

The CRyPTIC data was used to assess the prognostic sensitivity, Specificity, PPV, and NPV. Since the DL models were trained with TP Portals data, we used the independent dataset from CRyPTIC to assess these four values for prognostic potential of each mutation and all two-combinations of the 49 mutations.

## 3. Results

*CRyPTIC* data was mined to identify noncanonical mutation associated with INH resistance. To perform this task, we chose the subset of the 919 patients who were phenotypically susceptible to INH at baseline experienced the emergence of one of the three canonical mutations. In all 27 patients met these criteria. Table 1 depicts the classification of the set of 919 patients. Of the 919 patients, 27 did not have any of the three canonical mutations in their baseline sample but experienced the emergence of at least one of the three mutations during their treatment. We refer to this group as the “*Emerged*” Group. An additional 330 patients had at least one of the canonical mutations in their baseline sample. We refer to this group as the “Initial Resistance” group. Finally, in 562 patients the three canonical mutations never emerged during treatment. We refer to this group as the “Not Emerged” group. These were mostly phenotypically INH susceptible (INH^S^) patients.

**Table 1.**
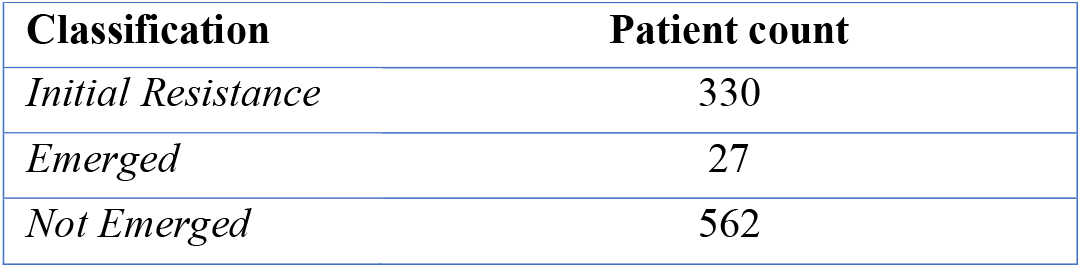
Classification of the 919 patients from CRyPTIC consortium based on the emergence of canonical mutations. “Initial” indicates a patient that had one of the three canonical mutations at the start of treatment. “Emerged” indicates that the patient had none of the canonical mutations but at least one of them emerged during treatment. “Not Emerged” are patients whose samples never harbored any of the canonical mutations.

| Classification | Patient count |
| --- | --- |
| *Initial Resistance* | 330 |
| *Emerged* | 27 |
| *Not Emerged* | 562 |

We used all isolates (73) collected from the 27 patients to identify mutations that were associated with emergence of the three canonical mutations. These would be all mutations that were observed in the nine identified genes in the 27 patients’ samples prior to the emergence of the canonical mutations. In total 49 mutations were identified through this exercise (Supplementary Table 1).

We then studied the predictive power of each of the 49 mutations through assessment of sensitivity, specificity, positive predictive value (PPV) and negative predictive value (NPV) in predicting the emergence of any of the three canonical mutations (*katG*315, *inhA*-15, *inhA*-8). The list of the top 12 mutations is depicted in Table 2.

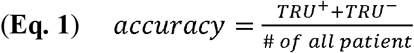

**Table 2.** List of 12 mutations that produced the best predictive models. The “(-)” sign indicates an inverse predictive relationship (the absence of the mutation is predictive of emergence of canonical mutations). PPV: Positive Predictive Value; NPV: Negative Predictive Value. AI model was trained with TB Portals data (Accuracy). Sensitivity, Specificity, PPV, and NPV was assessed using the CRyPTIC data.

| Mutation | AI Model Accuracy | Sensitivity | Specificity | PPV | NPV |
| --- | --- | --- | --- | --- | --- |
| *Rv1258c* 581 indel | 73.23% | 55.18% | 84.70% | 69.61% | 74.84% |
| *mshA* A187V | 71.82% | 45.51% | 82.59% | 62.31% | 70.56% |
| *ndhA* -72 indel | 63.55% | 12.92% | 95.38% | 63.89% | 63.40% |
| (-) *mshA* N111S | 62.67% | 4.78% | 90.94% | 25.00% | 60.16% |
| *inhA* S94A | 62.02% | 1.97% | 99.82% | 87.50% | 61.69% |
| *inhA* G3O | 61.81% | 3.09% | 98.76% | 61.11% | 61.71% |
| *katG* R463L | 61.67% | 74.44% | 31.26% | 40.64% | 65.92% |
| (-) *ahpC* G-88a | 61.67% | 11.20% | 66.90% | 17.70% | 54.26% |
| *fabG1* g-17t | 61.59% | 0.84% | 99.82% | 75.00% | 61.42% |
| *Rv2752c* L355L | 61.37% | 0.84% | 99.82% | 75.00% | 61.42% |
| *ndhA* G391R | 61.26% | 3.65% | 97.51% | 48.15% | 61.55% |
| *Rv2752c* A520A | 57.89% | 6.74% | 90.23% | 30.38% | 60.48% |

(INH<sup>S</sup>) patients.

### Machine Learning results

In this exercise 49 sets of models were constructed, one for each mutation (Supplementary Table 1) using the TB Portals data. The best 12 mutations for predicting the emergence of the three canonical mutations using DL and the model’s prediction accuracy (Eq. 1), for each of these mutations are also shown in Table 2. As can be seen, the best performing model used an indel (single-base insertion or deletion) at *Rv1258c* 581. Using this mutation, the model was able to reach a predictive accuracy of 69.33%. Mutations in the table are ordered based on the DL model accuracy as we believed it ranked the best predictive mutations the highest.

Because DL models can exploit inverse relationships for their predictions, they can exploit mutations whose presence is predictive of lack of emergence. The inverse relationships can be exploited for continued use of specific drugs in new regimens (when needed) to improve sterilization. To this point, it is important to note that two mutations (*mshA* N111S, *ahpC* G-88a) had an inverse relationship, meaning that their absence was predictive of emergence and their presence was predictive of non-emergence and potential continued sensitivity.

To further improve the prediction accuracy, we built models that used two mutations for their prediction. We exhaustively built models for every combination of two mutations from the list of 49. This produced 1,176 (_*49*_*C*_*2*_: 49 choose two) combinations and hence 1,176 sets of models. For each combination we trained 10 models.

The highest accuracy of all 1,225 (49 single mutation and 1,176 double mutation) DL model sets is shown in Figure 1. Table 3 shows the best accuracies achieved with the top eight two-mutation models. Table 3, unlike Table 2, is ranked based on specificity. We believe that specificity for the double mutation models is the most important feature since accuracy does not take into account the high false positive and false negative rates of these models. The highest prediction accuracy (77.33%) belonged to the model trained with the combination of two mutations: *ndhA* M325T and *Rv1258c* 581 indel. This model, however, has a low specificity at 66.25% due to its high rate of false positive calls. The second highest accuracy (75.00%) belonged to the combination of *ahpC* g-88a & *mshA* N111S mutations. This model also suffered from a low specificity (42.70%). Importantly, the best performing combinatorial model was *Rv1258c* 581 indel & *mshA* A187V with a specificity of 85.94% and sensitivity of 49.86%. While this sensitivity value is not the highest it is well within the range of what we expected (see discussions). Of interest is that since both *ahpc* g-88a and *mshA* N111S have an inverse relationship with emergence of the canonical mutations, the model that uses both of these mutations also uses the absence of these mutations as a signal of emergence.

**Table 3.**
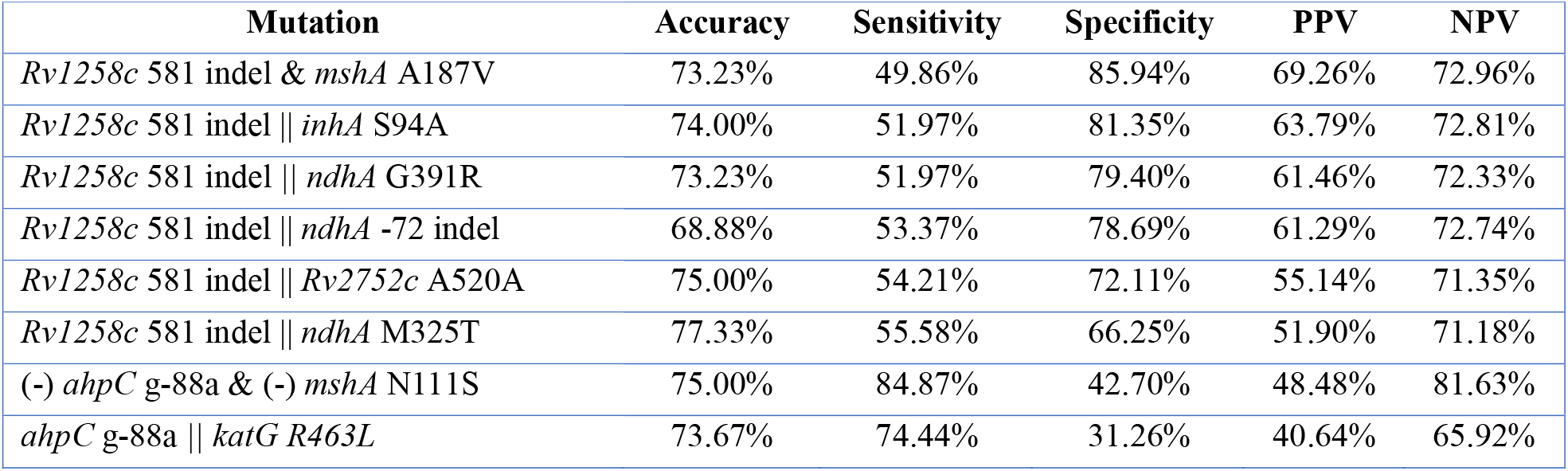
List of double mutations that produced the best five predictive models of the emergence. “(-)” indicates an inverse relationship with the emergence of canonical mutations. ‖ indicates the OR operator. I.e. the presence of either mutation is used for prediction. & indicates the AND operator. I.e. the presence of both mutations is used for prediction. AI models were trained with TB Portals data (Accuracy). Sensitivity, Specificity, PPV, and NPV was assessed using the CRyPTIC data.

**Figure 1.**
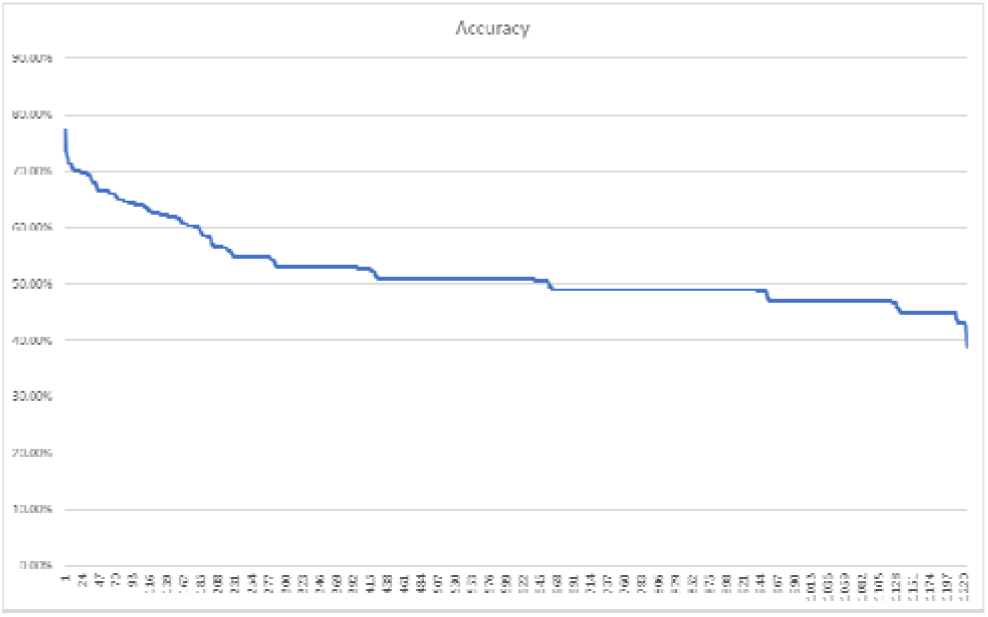
Deep Neural model accuracy in predicting the three canonical mutations (*katG*315, *inhA*-15, *inhA*-8) using at most two of the list of 49 candidate mutations (i.e. _*49*_*C*_*1*_ + _*49*_*C*_*2*_ = 1,225 combinations).

### Best predictive model

The selection of the best predictive model (*Rv1258c* 581 indel) is straight forward among the single mutation models as the highest accuracy belong to a model that also enjoyed the second highest sensitivity, highest NPV (74.84%), and an acceptable specificity (84.70%). A similar pattern follows for the second-best predictive mode (*mshA* A187V). However, this choice among the two-mutations model was not trivial as the model with highest accuracy (*Rv1258c* 581 indel || *ndhA* M325T) had an unacceptably high rate of false positive calls resulting causing its low specificity (66.25%). Alternatively, the model with highest sensitivity (*ahpC* g-88a & *mshA* N111S) suffered from lowest specificity and the model with highest specificity (*Rv1258* 581 indel & *mshA* A187V) suffered from the lowest sensitivity. In choosing the best model, we ranked higher specificity as more important than sensitivity, as long as sensitivity was not too low (see Discussions). To demonstrate the reasoning behind this choice, we present the performance of the two models with orthogonal performance characteristics against each other in Table 4. As it can be seen, the model with high sensitivity but low specificity is making many false positive calls (245 cases). Inversely, the model with high specificity and lower sensitivity is making some false negative calls (128 cases), albeit not as many as the false positive calls of the other model. Because the model with highest specificity (*Rv1258* 581 indel & *mshA* A187V) also had an the highest NPV and acceptable PPV and sensitivity, we chose this model to be the best performing double-mutation model.

**Table 4.** Performance correlation of two orthogonal models. Numbers in each box are the # of patients whose prediction by the two models fall into the category given by the column and row headers.

|  |  | *Rv1258c* 581 indel & *mshA* A187V |  |  |  |
| --- | --- | --- | --- | --- | --- |
|  |  | TRU+ | TRU- | FALSE+ | FALSE- |
| *ahpC* g-88a & *mshA* N111S | TRU+ | 175 | 0 | 0 | 128 |
|  | TRU- | 0 | 238 | 2 | 0 |
|  | FALSE+ | 0 | 245 | 77 | 0 |
|  | FALSE- | 3 | 0 | 0 | 51 |

Overall, Table 5 shows the best three predictive models. We picked the best performing model to be the single-mutation model using the mutation *Rv1258c* 581 indel to make its prediction. We chose this model over the double mutation model because of its similar specificity but slightly better sensitivity.

**Table 5.**
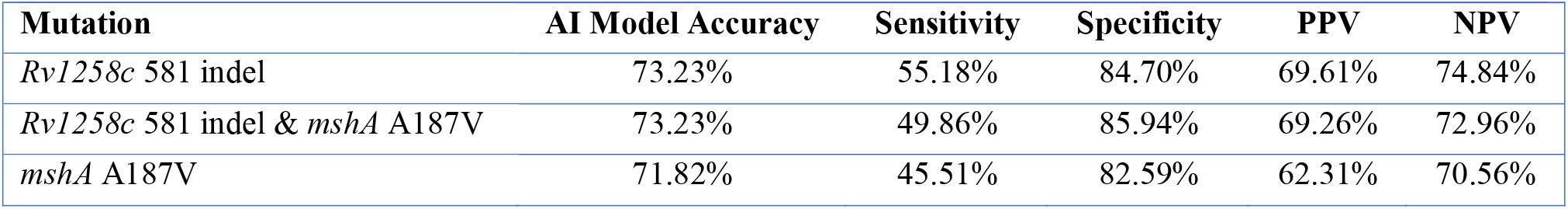
List of mutations that produced the best top predictive models of emergence of *katG*315, *inhA*-15, or *inhA*-8. PPV: Positive Predictive Value; NPV: Negative Predictive Value. AI models were trained with TB Portals data (Accuracy). Sensitivity, Specificity, PPV, and NPV was assessed using the CRyPTIC data.

## 4. Discussion

While new drugs infrequently are developed to combat TB, the emergence of resistance significantly reduces their shelf life, further reducing the appetite of the pharmaceutical companies for antibiotic drug development. As such, when resistance emerges, alternative treatment options are limited due to the limited drug choices. Prevention of emergence of resistance, therefore, must remain a priority for the medical research community. Since INH resistance often emerges first in a sequence of continued acquisition of additional resistance, prognosis of INH resistance carries a special significance in the battle against drug resistant tuberculosis with the hope that prevention of INH resistance will prevent or slow acquisition of resistance to other drugs as well.

In this manuscript we present an approach to identify and assess the prognostic potential of promising mutations for emergence of canonical mutations. Specifically, we present *Rv1258c* 581 indel and *mshA* A187V as the most promising precursor mutations for emergence of INH canonical resistance. The high specificity of our predictions (as high as 85.94%) demonstrates the potential of these mutations in predicting the emergence of canonical mutations. Furthermore, the existence of such precursor mutations indicates a stepwise molecular evolutionary trajectory towards high level isoniazid resistance at least for substantial percentage of resistance cases. This is a hypothesis that should be tested in animal models.

The combination of the two mutations, *Rv1258c* 581 indel and *mshA* A187V, was observed in 73% of the isolates collected from patients who experienced emergence of at least one of the three INH canonical mutations (*katG*315, *inhA*-15, and *inhA*-8) during treatment. In 89% of patients who had more than two-time course samples (i.e. had sufficient temporal sampling resolution) and experienced the emergence of the canonical mutations, at least one of the two precursor mutations was observed. The number of patients with sufficient temporal sampling resolution was small, even in the large CRyPTIC data set with over 12,000 samples. The great majority of patients only had one sample followed by the group of patients with two samples. This points to the need for a more systematic collection of samples that would consider their usability in studies such as this one. Upon collection of more time course samples, the study presented in this manuscript should be repeated to build more accurate models.

### Sensitivity and specificity

From the onset of the study, we anticipated low sensitivity but hoped for high specificity. We anticipated low sensitivity for a number of reasons. First, we aimed to predict specific resistance mutations (only three canonical mutations) from among tens of mutations that have been identified to cause INH resistance^17^. As such, we are only assessing a limited number of evolutionary trajectories to resistance. Furthermore, due to limited number of samples, we have only been able to expose precursor mutations indicative of some evolutionary trajectories toward the three canonical mutations. Due to the limited number of samples, we believed that this exercise would be unlikely to expose all evolutionary trajectories toward emergence of the three canonical mutations. Finally, since our entire sequencing data from both datasets are recorded on Illumina platforms, we had to exclude some genes such as *iniB* entirely from this study due to the impact of Illumina “blind spots” on proper sequencing of these genes.^20^ Among the genes impacted by these blind spots is also our top gene *Rv1258c*. While the region affected by the blind spot in this gene is very small and is not directly in position 581, because of the nature of the mutation (indel), the effect of other indel mutations in the gene that may be obscured by the blind spot may compromise the detection accuracy of the indel at position 581. For these reasons, among others, a lower sensitivity was expected. As such, reaching a sensitivity of over 55% was higher than our expectations indicating that perhaps the diversities of trajectories toward the canonical mutations are more limited than we expected. The very high specificity in combination with acceptable NPV and PPVs is promising. With more qualifying cases, and a more comprehensive set of mutations, these values are expected to further improve.

### *Rv1258c* 581 indel

Previous work has studied *Rv1258c* extensively. The consensus is that this gene is an efflux pump which also acts as a growth control factor^22^ through novel functions of oxidative stress and the VII secretion system *ESX-3* mediated iron metabolic pathway^23^. The pump is also implicated in resistance to a wide range of anti-TB drugs. Engineered strains with knocked out *Rv1258c* showed a slightly lower MIC for rifampin, ethambutol, ofloxacin, amikacin, capreomycin and streptomycin.^22^ Mutations in the gene have also been reported to cause resistance to ethambutol and capreomycin^22^, and to pyrazinamide, isoniazid, and streptomycin^24^. Li et. al. reported MIC values for strains with the same insertion as the one reported here (*Rv1258c* 581 T Ins G).^25^ Three strains with this mutation were reported to have an MIC value of 4. One also had *katG* Trp351Gly mutation, the second had the addition mutation *ahpC* -641 G>A, and the third had the additional mutation *ahpC* -74 G>A. While mutations other than our reported 581 T Ins G in the gene have been reported to cause high level of resistance in H37Ra^24^, we hypothesize that 581 T Ins G may result in a slightly elevated levels of drug efflux, and hence tolerance to the drug, buying time for the canonical mutations to emerge and cause high levels of resistance. If this hypothesis is proven to be correct, prevention of high-level INH resistance is possible for cases in which *Rv1258c* 581 indel is observed. This hypothesis can be tested with a mutagenesis confirmation exercise.

### *mshA* A187V

The gene *mshA*, in particular, the mutation (*mshA* A187V) reported here is less studied. The gene encodes for a glycosyltransferase.^26^ Buchmeier et. al. concluded that the gene is essential for growth.^26^ Barth et. al. determined that the expression of the gene is increased significantly in response to oxidative and copper stress.^27^ Xu et. al. determined that *mshA* deletion results in isoniazid resistance.^28^ Jagielski et. al. concluded that the mutation identified in this manuscript (*mshA* A187V) did not cause resistance since they observed the mutation in four resistant (three MDR and one mono INH^R^) and one INH^S^ isolate.^29^ It is possible that this mutation also causes low level resistance below the critical concentration and hence by itself would not reduce susceptibility enough to pass the critical concentration and therefore isolates only harboring this mutations would be classified as susceptible. This hypothesis can also be tested through a mutagenesis experiment similar to the previous mutation. If this hypothesis proves to be incorrect, the relationship with INH resistance will require a broader set of hypotheses and experiments.

### *Rv1258c* 581 indel and *mshA* A187V epistasis

While the two mutations together provide notable predictive value for emergence of canonical isoniazid mutations, it is difficult to see how the two can operate in any epistatic form since the two genes are almost one million bases away. As such, any such interaction may be conceivable from a functional pathway perspective. As far as we can determine, these two genes are not known to be part of the same pathway despite their similar roles to some stressors. We hypothesize that this is likely independent emerging responses of the pathogen to drug pressure through two different pathways that respond to similar external pressures. The two mutations identified here may enhance such a response—a hypothesis that should be tested. The coexistence of the two mutations may mean a more potent tolerance to INH pressure and hence likely survival for longer periods of time and eventual emergence of mechanisms that provide higher levels of resistance. The fact that we have observed only one of the two mutations in some patients provides some support to this hypothesis. The hypothesis can be tested by introducing the mutations into a wildtype strain prior to introduction of INH.

For clinical applications, the prognostic significance of the two mutations provides hope that prediction of at least canonical mutations may be possible for most cases.

In this project, we only considered the prognostic significance of up to two mutations at a time. This was solely done in consideration of the exponentially increasing computational complexity with increasing combinatorial complexity (NP-complete class of problems). As such, this project only exposes the most prominent prognostic markers for the most prominent evolutionary trajectories to emergence of the three canonical mutations. Given these limitations, the prognostic accuracy of 73% with specificity of over 85% is quite promising. Further clinical data collection and experimentation that removes some of these limitations, such as consideration of at least three mutations at a time is expected to improve the sensitivity to levels acceptable for clinical implementation.

## 5. Data Availability

All data used in this project is publicly available through two databases: TB Portals and the CRyPTIC consortium. All data produced in the present work are contained in the manuscript.

